# Predictions of COVID-19 dynamics in the UK: short-term forecasting and analysis of potential exit strategies

**DOI:** 10.1101/2020.05.10.20083683

**Authors:** Matt J. Keeling, Edward M. Hill, Erin E. Gorsich, Bridget Penman, Glen Guyver-Fletcher, Alex Holmes, Trystan Leng, Hector McKimm, Massimiliano Tamborrino, Louise Dyson, Michael J. Tildesley

**Author notes:** These authors contributed equally to this work (Edward M. Hill, Louise Dyson, Michael J. Tildesley).

## Abstract

**Background:** Efforts to suppress transmission of SARS-CoV-2 in the UK have seen non-pharmaceutical interventions being invoked. The most severe measures to date include all restaurants, pubs and cafes being ordered to close on 20th March, followed by a “stay at home” order on the 23rd March and the closure of all non-essential retail outlets for an indefinite period. Government agencies are presently analysing how best to develop an exit strategy from these measures and to determine how the epidemic may progress once measures are lifted. Mathematical models are currently providing short and long term forecasts regarding the future course of the COVID-19 outbreak in the UK to support evidence-based policymaking.

**Methods:** We present a deterministic, age-structured transmission model that uses real-time data on confirmed cases requiring hospital care and mortality to provide up-to-date predictions on epidemic spread in ten regions of the UK. The model captures a range of age-dependent heterogeneities, reduced transmission from asymptomatic infections and produces a good fit to the key epidemic features over time. We simulated a suite of scenarios to assess the impact of differing approaches to relaxing social distancing measures from 7th May 2020 on the estimated number of patients requiring inpatient and critical care treatment, and deaths. With regard to future epidemic outcomes, we investigated the impact of reducing compliance, ongoing shielding of elder age groups, reapplying stringent social distancing measures using region based triggers and the role of asymptomatic transmission.

**Findings:** We find that significant relaxation of social distancing measures from 7th May onwards can lead to a rapid resurgence of COVID-19 disease and the health system being quickly overwhelmed by a sizeable, second epidemic wave. In all considered age-shielding based strategies, we projected serious demand on critical care resources during the course of the pandemic. The reintroduction and release of strict measures on a regional basis, based on ICU bed occupancy, results in a long epidemic tail, until the second half of 2021, but ensures that the health service is protected by reintroducing social distancing measures for all individuals in a region when required.

**Discussion:** Our work confirms the effectiveness of stringent non-pharmaceutical measures in March 2020 to suppress the epidemic. It also provides strong evidence to support the need for a cautious, measured approach to relaxation of lockdown measures, to protect the most vulnerable members of society and support the health service through subduing demand on hospital beds, in particular bed occupancy in intensive care units.

## Introduction

In late 2019, accounts emerged from Wuhan city in China of a virus of unknown origin that was leading to a cluster of pneumonia cases [1]. The virus was identified as a novel strain of coronavirus on 7th January 2020 [2] and the first known death as a result of the disease occurred two days later [1]. Over the next few days, cases were reported in several other cities in China and in other countries around the world including South Korea, Japan and the United States of America. On 23rd January, the Chinese government issued an order for Wuhan city to enter “lockdown”, whereby all public transport was suspended and residents were not allowed to leave the city. Over the next 24 hours, these measures were extended to all the major cities in Hubei province in an attempt to prevent further spread of disease.

Whilst the introduction of these severe social distancing measures began to have an effect upon reducing the growth rate of cases in Wuhan [3–5], reported cases outside China continued to grow and by late February the virus, now designated by the World Health Organisation as SARS-CoV-2, and the disease it causes as coronavirus disease 2019 (COVID-19), had spread to Europe, with a growing number of cases being reported in northern Italy [6]. As more countries in Europe and around the world started to experience a dramatic rise in cases, similar measures were put in place in an effort to protect the most vulnerable members of society and to ensure that health services capacities were not exceeded [6, 7].

In the UK, the first cases of COVID-19 were reported on 31st January 2020, in the city of York in the north of England. In the early stages of the UK outbreak, the government focused on a strategy of containment, to reduce the likelihood of large-scale within-country transmission occurring. This strategy involved rapid identification and isolation of infected individuals, achieved through contact tracing and testing of suspect cases. However, by early March it was evident that sustained community transmission was occurring and there was a growing concern that a large epidemic could rapidly overwhelm the health service, resulting in a significant number of deaths. This led to the government considering the introduction of a range of social distancing measures in order to slow the growth of the outbreak, thus delaying and flattening the epidemic peak and reducing the risk of exceeding hospital capacities owing to an influx of COVID-19 patients. On 12th March, the UK officially entered the “delay” phase, with the government declaring that all individuals with a cough or fever should self-isolate for a period of seven days. Over the following days, several major sporting events were cancelled and the public was advised to avoid all non-essential travel and contact with others. With daily cases and deaths continuing to rise, the government introduced its most severe measures: all restaurants, pubs and cafes were ordered to close on 20th March; schools were also ordered to close on 20th March except for the children of key workers; finally a “stay at home” order was issued on the evening of 23rd March together with the closure of all non-essential retail outlets for an indefinite period. By this time the reported number of deaths in the UK had reached 335.

This paper was originally written in April 2020, and throughout we use the epidemiological data up to 21st April in all figures [8]. The data and science surrounding the SARS-Cov-2 infection is fast moving, so much so that publications can rarely keep pace. We therefore intend this paper to be a record of the state of our predictive modelling in mid-April, just after the peak of the first wave, although we comment more fully in the discussion about later improvements to the model formulation and the implications of the results for controlling the later second wave.

Even at the peak of the UK epidemic, it was clear that the stringent lockdown rules imposed could not continue indefinitely. It was apparent that epidemiological modelling was a vital tool for analysing potential “exit strategies”, which could allow some relaxation of social distancing measures, whilst minimising the future impact of the disease on the health service. At the time epidemiologists were critically aware that, should measures be relaxed too rapidly when there were still sufficient susceptible individuals in the population, there was a high risk of a second infection wave that could once again threaten to overwhelm health services. The increasing cases, hospitalisations and deaths observed in the UK (and elsewhere in the world) during September and October of 2020 confirms our earlier predictions, and strengthens the need for a robust and managed exit strategy.

In this paper, we present a novel compartmental mathematical model of SARS-CoV-2 transmission tailored to attributes notable to COVID infections, including household saturation of transmission, household quarantining and age-dependent detectability and transmission of SARS-CoV-2. The model uses real-time data on confirmed cases requiring hospital care and mortality to provide short and long term forecasts on epidemic spread in ten regions of the UK. We investigate how compliance with social distancing affects future epidemic outcomes. We compare and contrast different exit strategies, namely: relaxing social distancing by age group, or the regional lifting and imposition of restrictions according to healthcare system capacity. Finally, we explore the sensitivity of our conclusions to a key biological aspect of SARS-CoV2 which remains unknown: whether different age groups differ in their core susceptibility to infection, or their likelihood of displaying symptoms.

## Methods

### Transmission model

Here, we describe a compartmental model that has been developed to simulate the spread of SARS-CoV-2 virus (resulting in cases of COVID-19) in the UK population. In the ongoing outbreak in the UK, cases of COVID-19 are confirmed based upon testing, with priority for testing throughout the majority of the initial wave given to patients requiring critical care in hospitals [9] - generating biases and under-reporting. There is evidence to suggest that a significant proportion of individuals who are infected may be asymptomatic or have only mild symptoms [10, 11]. These asymptomatic individuals are still able to transmit infection [12], though it remains unclear whether they do so at a reduced level. Our modelling approach has consequently been designed to elucidate the interplay between symptoms (and hence detection) and transmission of SARS-CoV-2.

We developed a deterministic, age-structured compartmental model, stratified into five-year age bands. Inclusion of age-structure within the model was considered critically important given the greater number of cases, hospitalisations and deaths amongst older age-groups. Transmission was governed through age-dependent mixing matrices based on UK social mixing patterns [13, 14]. The population was further stratified according to current disease status, following a susceptible-exposed-infectious-recovered (SEIR) paradigm, as well as differentiating by symptoms, quarantining and household status (Fig. 1). Susceptibles (*S*) infected by SARS-CoV-2 entered a latent state (*E*) before becoming infectious. Given that only a proportion of individuals who are infected are tested and subsequently identified, the infectious class in our model was partitioned into symptomatic (and hence potentially detectable), *D*, and asymptomatic (and likely to remain undetected) infections, *U*. We assumed both susceptibility and disease detection were dependent upon age, although the partitioning between these two components is largely indeterminable (additional details are given in Table 1 and Supporting Text S1). We modelled the UK population aggregated to ten regions (Wales, Scotland, Northern Ireland, East of England, London, Midlands, North East and Yorkshire, North West England, South East England, South West England), with each region modelled independently (i.e. we assumed no interactions occurred between regions).

**Table 1:** Key model parameters.

| Parameter | Description | Value | Source |
| --- | --- | --- | --- |
| $\beta_{ba}$ | Age-dependent transmission from age group $b$ towards age group $a$, split into household, school, work and other | | POLYMOD matrices [14] |
| $\epsilon$ | Rate of progression to infectious disease ($1/\epsilon$ is the duration in the exposed class) | $\sim 0.2$ | Fitted as part of MCMC process |
| $\gamma$ | Recovery rate, changes with $\tau$, the relative level of transmission from undetected asymptomatics compared to detected symptomatics | $\sim 0.5$ | Fitted from early age-stratified UK case data |
| $\alpha$ | Scales whether age-structure case reports are based on age-dependent symptoms ($\alpha = 0$) or age-dependent susceptibility ($\alpha = 1$) | $0 - 1$ | Fitted as part of MCMC process |
| $\tau$ | Relative level of transmission from asymptomatic compared to symptomatic infection | $0 - 0.5$ | Fitted as part of MCMC process |
| $d_a$ | Age-dependent probability of displaying symptoms (and hence being detected), changes with $\alpha$ and $\tau$ | | Fitted from early age-stratified UK case data |
| $\sigma_a$ | Age-dependent susceptibility, changes with $\alpha$ and $\tau$ | | Fitted from early age-stratified UK case data |
| $\phi$ | Impact of adherence with restrictions | $0 - 1$ | Fitted as part of MCMC process or varied according to scenario |
| $H$ | Household quarantine proportion | $0 - 1$ | Can be varied according to scenario |
| $N_a$ | Population size of a given age | By region | ONS |

**Fig. 1:**
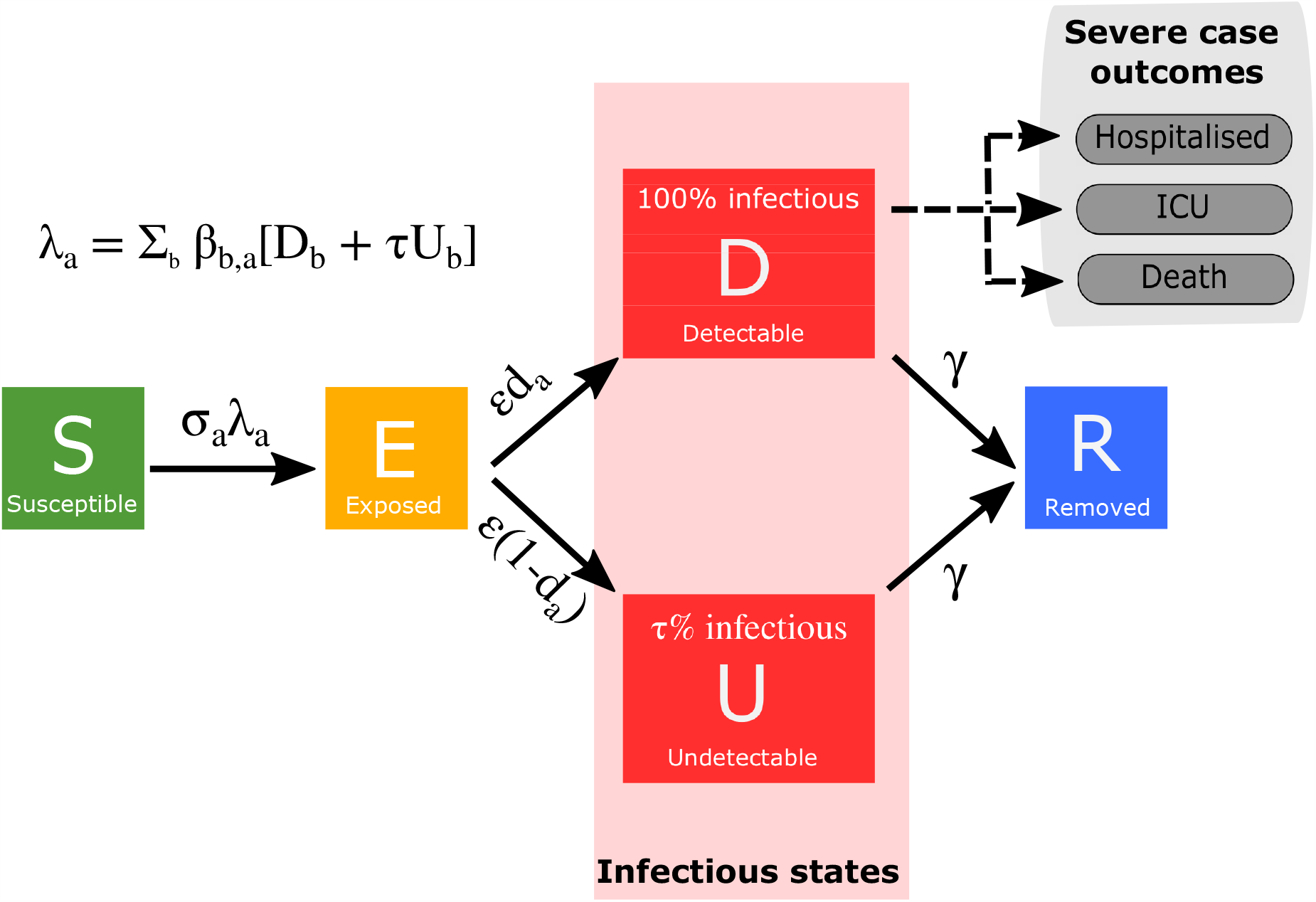
Disease states and transitions. We stratified the population into susceptible, exposed, detectable infectious, undetectable infectious, and removed states. Solid lines correspond to disease state transitions, with dashed lines representing mapping from detectable cases to severe clinical cases that require hospital treatment, critical care (ICU), or result in death. The model was partitioned into five-year age bands. See Table 1 for a listing of model parameters. Note, we have not included quarantining and household status on this depiction of the system.

A drawback of the standard SEIR ordinary differential equation (ODE) formation in which all individuals mix randomly in the population is that it cannot readily account for the isolation of households. For example, if all transmission outside the household is set to zero in a standard ODE model, then an outbreak can still occur as within-household transmission allows infection between age-groups and does not account for local depletion of susceptibles within the household environment. We addressed this limitation by extending the standard SEIR models such that first infections within a household (*E*^*F*^, *D*^*F*^, *U* ^*F*^) are treated differently from subsequent infections (*E*^*S*^, *D*^*S*^, *U* ^*S*^). To account for the depletion of susceptibles in the household, we made the approximation that all within household transmission was generated by the first infection within the household (for further details, see Supporting Text S1).

### Case severity parameterisation

The model is concerned with epidemiological processes and so predicts the number of symptomatic and asymptomatic infections on each day. However, in order to provide evidence regarding the future impact of the outbreak in the UK, it is crucial to be able to predict the number of severe cases that may require hospital or critical care. We utilised two processes in order to estimate hospitalisation rates: (i) we estimated the proportion of clinical cases in each age group that would require hospitalisation by comparing the age distribution of hospital admissions to the age structure of early detected cases — assuming these detected cases were an unbiased sample of symptomatic individuals; (ii) we used age independent distributions to determine the time between onset of symptoms and hospitalisation. A similar process was repeated for admission into intensive care units. Both of these distributions were drawn from the COVID-19 Hospitalisation in England Surveillance System (CHESS) data set that collects detailed data on patients infected with COVID-19 [15] (for further information, see Supporting Text S2).

Information on the distributions of length of stay in both intensive care units (ICUs) and hospital was used to translate admissions into bed occupancy — which adds a further delay between the epidemiological dynamics and quantities of interest.

In terms of matching the available data and quantities of interest, we also use the prediction of symptomatic infections to drive the estimated daily number of deaths within hospitals. The risk of death is again captured with an age-dependent probability, while the distribution of delays between hospital admission and death is assumed to be age-independent. These two quantities are determined from the Public Health England (PHE) death records.

### Model fitting

We fit the model framework to each of the ten UK regions independently, on a region-by-region basis, to four timeseries: (i) new hospitalisations; (ii) hospital bed occupancy; (iii) ICU bed occupancy; (iv) daily deaths (using data on the recorded date of death, where-ever possible).

The relative transmission rate from asymptomatic cases (*τ*) and the scaling of whether age-structure case reports were based on age-dependent susceptibility or age-dependent symptoms (*α*) were treated as free parameters. These allowed us to define an age-dependent susceptibility (*σ*_*a*_) and an age-dependent probabilities of displaying symptoms (*d*_*a*_), through the next-generation relationship:

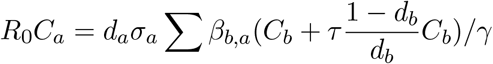

which linked observed cases in the next generation to the number of observed and unobserved infections in the previous week. By assuming that the two age-dependent probabilities were linked by:

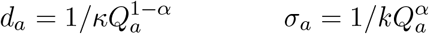

we were able to obtain the probabilities that were consistent with the age-distribution of observed cases, and the required basic reproductive ratio *R*_0_ (see Supporting Text S1 for further details and [16] for more information on the inference scheme).

We performed parameter inference using the Metropolis-Hastings algorithm, computing likelihoods assuming the daily count data for the four quantities to be independently drawn from Poisson distributions, with a mean equal to the value derived from the model [16]. After a burn-in of 250,000 iterations, the algorithm was run for a further 250,000 iterations. We thinned the generated parameter sets by a factor of 100, giving 2,500 parameter sets representing samples from the parameter posterior distributions. Example posterior distributions for key parameters are given in Supporting Text S3; in all cases we use relatively uninformative priors and observe substantial departure from the prior distributions.

### Modelling intervention scenarios

In order to capture the impact of social distancing measures that were introduced in the UK (on 23rd March) to reduce transmission, we scaled down the mixing matrices associated with schools, work and other activities while increasing the within household transmission matrix (see Supporting Text S4). This approach allowed us to flexibly vary the effectiveness of different social distancing measures and investigate the impact of compliance with social distancing (*ϕ*) upon the future spread of disease. We considered a range of compliance levels, scaling from zero (no-compliance) to one (maximal compliance), as well as inferring the compliance parameter from the available data (*ϕ* = 0.53(0.36 − 0.70) across all regions).

Another prominent intervention measure to reduce the spread of infection has been household quarantining, whereby an entire household is instructed to self-quarantine when any member of that household starts to show symptoms of infection. To incorporate household quarantining measures into the ODE formulation, we added a quarantined class into our model, whereby a fraction (*H*) of the first detectable infection in any household (and therefore by definition a symptomatic case) is quarantined as are all their subsequent household infections. Accounting for the effect of household saturation also ensures that subsequent household infections do not contribute to further transmission. For a complete description of the model equations, see Supporting Text S1.

We used this model framework to perform a series of analyses assessing the impact of social distancing strategies on the future spread of infection. Unless otherwise stated, all interventions shown represent the mean dynamics from the posterior parameters inferred by a Monte Carlo Markov Chain (MCMC) fitting scheme, rather than the combination of the mean plus the sampling distribution; where practical credible intervals are also shown.

### Short term projections under current lockdown measures

To provide a baseline for comparison of our intervention scenarios, we initially simulated our model to investigate the impact of the current intervention policies, continuing from their introduction on 23rd March 2020. We simulated the model from 1st March 2020 to 30th April 2020 and compared the results to a scenario where no lockdown measures were ever introduced. To quantify prediction uncertainty, a total of 200 simulations were run for each scenario (lockdown activated or no lockdown imposed) using distinct parameter sets produced by the MCMC procedure, representing samples from the posterior parameter distributions. We focused our attention on estimates of deaths as well as hospitalisation and ICU bed occupancy, as key public-health considerations.

### Age-independent relaxation of lockdown measures

To investigate the longer term impact of the epidemic, we explored several scenarios in which control measures are relaxed on 7th May. The first scenario investigated a policy whereby social distancing measures were relaxed on 7th May for all individuals, regardless of age. To reflect the uncertainty in the degree of relaxation of the lockdown at this point, we varied our social distancing compliance parameter (*ϕ* = 0, 0.25, 0.5, 0.75, 1), which allowed us to consider how the epidemic trajectory may be affected for a range of relaxation policies. In these simulations we assumed that any remaining social distancing measures were fully removed at the end of 2020.

### Age-dependent relaxation of lockdown measures

Given the far higher fatality levels observed in the elderly, we next investigated policies imposing age-dependence upon the relaxation criteria. Specifically, we allowed all social distancing measures to be lifted from 7th May for any individual below a certain age (this age threshold was varied between 45 to 75 year old). For those above the age threshold, we assumed that social distancing measures remained in place until the end of 2020. Simulations were then run to the end of 2021, to capture any subsequent waves of infection. For each age threshold under consideration, we again considered the cumulative deaths, as well as cumulative hospital and ICU bed occupancy. We differentiated between these health impacts that occurred when age-specific restrictions were in place and when all restrictions were lifted. We also focused on the number of days in which ICU bed occupancy exceeded 4,000, as a measure of the immediate severity of the outbreak and the pressure on the health services.

### Full relaxation of lockdown measures with region-based reintroduction

Our penultimate set of simulations considered an adaptive intervention strategy, whereby lockdown measures were fully relaxed on 7th May, but then reintroduced when occupancy of intensive care units exceeded a given capacity and relaxed again when ICU occupancy declines. To account for regional variation in the outbreak and local hospital capacities, we assumed that control measures would operate locally (using the ten regions). We therefore used a pro-rata threshold, which equated to 3,000 occupied beds on a nationwide scale, as a trigger for reintroducing or relaxing controls (see Table S2). Given the sizeable delay between the implementation of controls and their effects on ICU occupancy, the dynamics only predicted a low number of switches between control and relaxation. We gathered regional predictions of daily deaths, ICU bed occupancy and hospital bed occupancy, with simulations run to the end of 2023.

### Sensitivity analysis

When evaluating the impact of lockdown measures, we are reliant upon recorded data on confirmed cases, hospital admissions and ICU occupancy in order to infer parameters of our model. However, there is still ongoing uncertainty in the relative level of transmission from asymptomatic individuals (*τ*) and the mechanisms driving age-specific detection rates (*α*). A range of *α* and *τ* parameter values are all able to generate predictions that closely match the available data. We therefore carried out a sensitivity analysis to these two parameters, investigating the impact of applying lockdown measures for specific age groups, as these parameters vary. We allowed *τ*, the relative level of transmission from asymptomatic individuals, to vary between 0 and 0.5; while *α* varied between 0 and 1. For large *α*, higher proportions of confirmed cases in a particular age group is as a result of greater susceptibility; whereas low vales of *α* indicate that a higher proportions of confirmed cases is due to greater severity of symptoms. This key parameter interacts with the relative transmission from asymptomatic infection (*τ*), although *τ* plays a minimal role when *α* is small. To assess the impact of these parameters on the effectiveness of lockdown measures, we computed the early epidemic growth rate under restrictions that target four specific age-groupings: (i) pre-school children under 5 (PS), (ii) school-aged children and young adults, 5-20 (S), (iii) adults between the age of 20 and 70 (A) and (iv) the elderly over 70 (E).

## Results

### Reductions in clinical case burden under current lockdown measures versus no intervention

Our model predicts that, should the current lockdown policies be continued, the number of daily deaths would peak in April across all regions before starting to decline (Fig. 2). England and Wales are found to be most severely affected, with the highest number of predicted deaths per capita, whilst we predict a lower number of deaths per capita in Scotland and Northern Ireland (noting that though our regional model fits generally had strong correspondence with the data, the fit to Scotland was weaker). All English regions show similar behaviour, other than the South East and South West, where we predict a lower number of deaths (Fig. 2). We observe similar behaviour in the levels of hospital and intensive care unit occupancy throughout this period (Fig. S2 and Fig. S4). Our model predicts that, under continued total lockdown, the average total deaths would be approximately 39,000 (Table 2).

**Table 2:** Summary of Model Outputs. *=epidemic would continue.

| Control | Time frame | Total deaths<br>(thousands) | Total QALY loss<br>(thousands) | ICU occupancy<br>(thousand bed days) | Hospital occupancy<br>(thousand bed days) | Lockdown scale<br>(days / population size) |
| --- | --- | --- | --- | --- | --- | --- |
| No Control | 1/1/2020 -<br>1/1/2021 | 201<br>(145-238) | 1997<br>(1140-2658) | 476<br>(204-671) | 3000<br>(1566-4161) | 0 |
| Observed lockdown* | 1/1/2020 -<br>1/1/2021 | 39<br>(36-42) | 315<br>(288-346) | 69<br>(62-75) | 462<br>(422-501) | 285 |
| Observed lockdown until<br>1/1/2021 then no control | 1/1/2020 -<br>1/1/2022 | 198<br>(131-234) | 1976<br>(974-2589) | 477<br>(180-657) | 2966<br>(1389-4009) | 285 |
| Regional switching<br>based on ICU occupancy | 1/1/2020 -<br>1/1/2023 | 155<br>(106-187) | 1352<br>(742-1817) | 280<br>(122-416) | 1933<br>(1029-2642) | 242<br>(121-347) |
| Lockdown of > 60s<br>until 2021 | 1/1/2020 -<br>1/1/2022 | 138<br>(111-165) | 1245<br>(721-1692) | 368<br>(151-530) | 1677<br>(886-2249) | 132 |

**Fig. 2:**
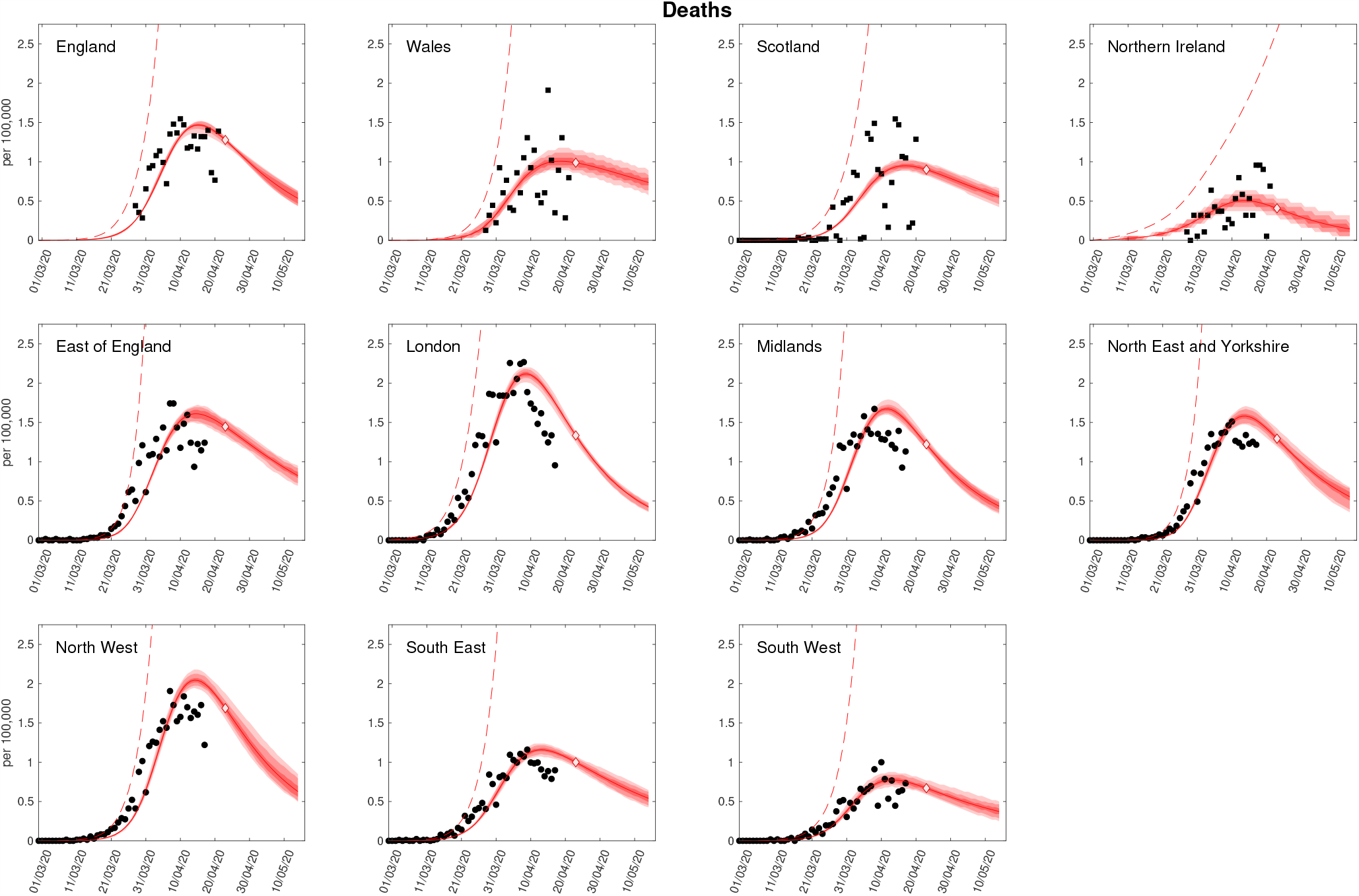
Regional projections for deaths per 100,000 with and without imposition of lockdown. In each panel: filled markers correspond to observed data (squares are for deaths by date of reporting, circles are for when date of death is available), solid lines correspond to the mean outbreak over a sample of posterior parameters; shaded regions depict prediction intervals, with darker shading representing stricter confidence (dark shading - 50%, moderate shading - 90%, light shading - 99%); red dashed lines illustrate the mean projected trajectory had no lockdown measures being introduced. We stress that the sample distribution around the expected value is not included in these plots, but would significantly increase the width of the distributions shown. (Prediction were produced on 23rd April, using data up to 21st April).

The fit to the available data is imperfect, which may be due to multiple factors. The data for the individual nations (top row) is by date of reported death, which introduces a number of reporting delays into the system. In addition, we are striving for a model that matches death, hospitalisations, hospital occupancy and ICU bed occupancy - the fits therefore represent a balance of fitting to all four measures (see the Supporting Information). Given the far greater numbers that are hospitalised, we find that these dominate the fitting procedure.

If the epidemic in the UK had been allowed to progress with no introduction of lockdown measures, our model predicts that the epidemic would have continued to grow throughout April, with deaths exceeding 200,000 by the end of 2021 (Table 2). This provides strong evidence to support the necessity of the social distancing measures that were introduced in order to reduce the growth rate of the epidemic and ensure that the health service was not overwhelmed with admissions.

### Measured age-independent relaxation protocols to reduce health system burden

Evaluating a policy whereby social distancing measures were relaxed to different degrees from 7th May for all individuals, we found that for a significant relaxation of lockdown the epidemic rapidly resurges with a peak in daily deaths of over 4,000 occurring in late June (Fig. 3, top panel). We project intensive care unit occupancy to near 10,000 by the end of June (Fig. 3, second panel), implying that significant release of lockdown measures would not be advisable. For more measured relaxation protocols, we found that, whilst there may be a slight resurgence in cases in the short term, hospital and ICU occupancy remained within capacity. However, whilst the forecasts from these simulations suggest that keeping most lockdown measures in place can have a positive impact upon reducing cases and deaths in the short-term, we note that, when lockdown measures are subsequently released in 2021 a large second infection wave is predicted. These results imply that should the outbreak have to be contained by non-pharmaceutical interventions alone, then a second wave of infections is somewhat inevitable as isolation measures are reduced. Of the scenarios investigated here, intermediate levels of relaxation (*ϕ* = 0.5) until 2021 followed by complete cessation of lockdown generates the least deaths (approximately 152,000 over both years).

**Fig. 3:**
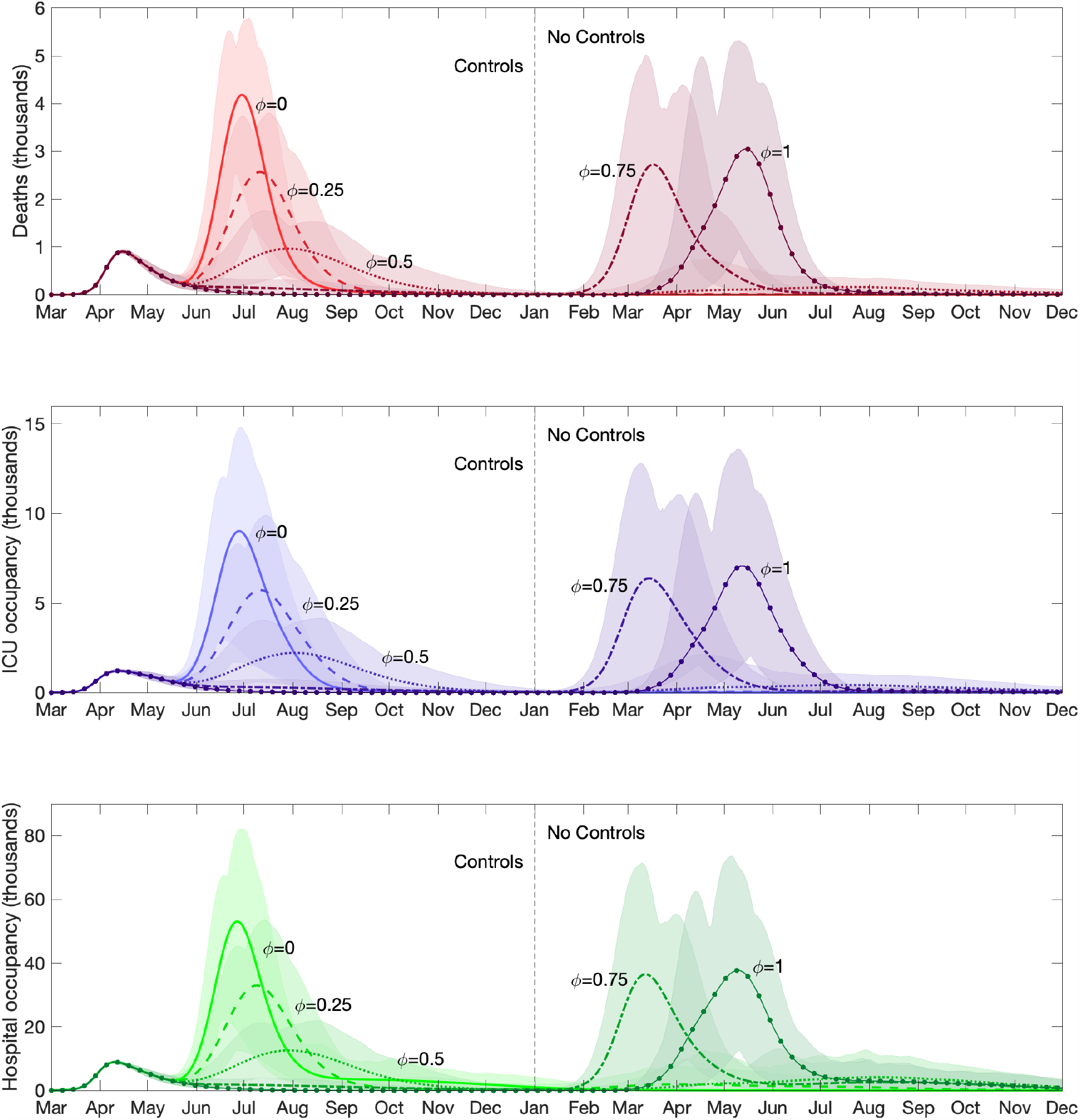
Clinical case projections for differing strengths of relaxing lockdown measures, *ϕ*. We assume social distancing measures were relaxed on 7th May for all individuals. The different line types (and shades) correspond to the dynamics using differing levels of relaxation (*ϕ* = 0, 0.25, 0.5, 0.75, 1), with *ϕ* = 0 corresponding to a total removal of social distancing measures, and *ϕ* = 1 representing a continuation of lockdown measures until 1st January 2021. Shaded regions represent the 95% posterior prediction intervals. We display daily counts of **(Row one)** deaths; **(Row two)** ICU occupancy; **(Row three)** hospital occupancy. At the start of 2021, all remaining social distancing measures are removed (the “no control” phase).

### Assessment of age-based shielding strategies

We next analysed the lifting of social distancing measures on 7th May for all individuals below a certain age, with social distancing measures remaining in place for the remainder of the population until the end of 2020. We observe that continuing lockdown for anyone over the age of 45 for the duration of 2020 results in the lowest number of deaths and number of admissions into hospital and ICU wards during that year (Fig. 4, first column). However, upon release of these lockdown measures we observed a significant second wave in 2021 as a substantial number in the over 45 age group were susceptible allowing a new outbreak (Fig. 4, second column). When isolation is only in place for older age groups (for example the over 70s), a large initial wave of infection occurs during 2020, but a subsequent secondary wave is not observed. Considering the combined impact from 2020-2021, we find that a strategy of continuing lockdown measures for anyone over the age of 65 minimises the total number of deaths, while hospital and ICU occupancy is minimised by continuing lockdown for anyone over the age of 60 although the overall effect of this is marginal (Fig. 4, third column). We predict that continuing lockdown for the over 60s throughout 2020 whilst relaxing measures of the remainder of the population results in, on average, 138,000 deaths by the end of 2021 (Table 2). Finally, we note that the total number of days for which ICU bed occupancy exceeds 4,000 increases with the age-threshold; this implies that while the elderly are the most vulnerable and the most likely to need critical care, an uncontrolled outbreak in the younger population can still place severe demands upon the health service (Fig. 4, third row).

**Fig. 4:**
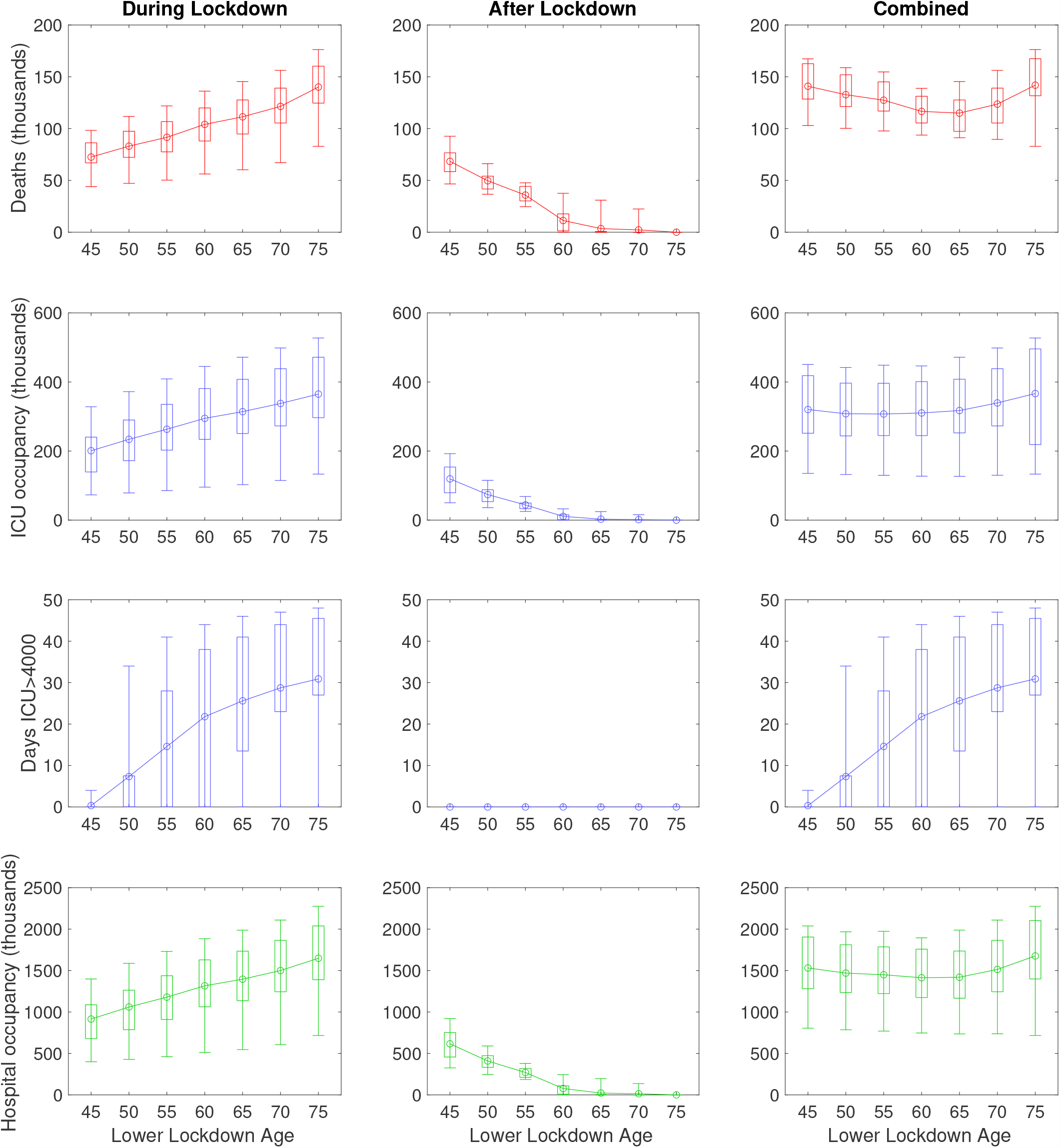
Impact of age-based shielding strategies on outbreak burden. In these simulations, social distancing measures were lifted on 7th May for all individuals below an age threshold, with social distancing measures remaining in place for the remainder of the population until the end of 2020. No interventions were applied post-lockdown release, with simulations continued until the end of 2022. Box plots for each statistic give median values (circles), interquartile range (box) and 95th percentiles (whiskers). Solid lines depict the profile of median estimates across age threshold space. The following statistics were computed for the period 23rd March 2020 to the end of 2021: **(Row one)** cumulative deaths; **(Row two)** cumulative ICU bed occupancy; **(Row three)** amount of days ICU occupancy exceeded 4000; **(Row four)** cumulative hospital bed occupancy. We stratify the outputs occurring across the considered time horizon in three ways: **(Column one)** during lockdown; **(Column two)** after lockdown; **(Column three)** combined (entire time horizon).

### Utility of reintroducing lockdown measures regionally with ICU occupancy triggers

Relaxing the lockdown from 7th May allows subsequent secondary waves of infection to begin, but a local increase in ICU occupancy triggers the reintroduction of social distancing measures on a region-by-region basis (Fig. 5). This results in multiple regional waves of infection, gradually becoming smaller and more asynchronous over time. The consequence of this adaptive strategy is that the total number of deaths and confirmed cases gradually reduce over a long period of time (Fig. 5, top and bottom panels), with the epidemic reaching low levels in mid 2021. As a result this policy balances the overall demand on the health services against the need to exit the epidemic without a substantive second wave.

**Fig. 5:**
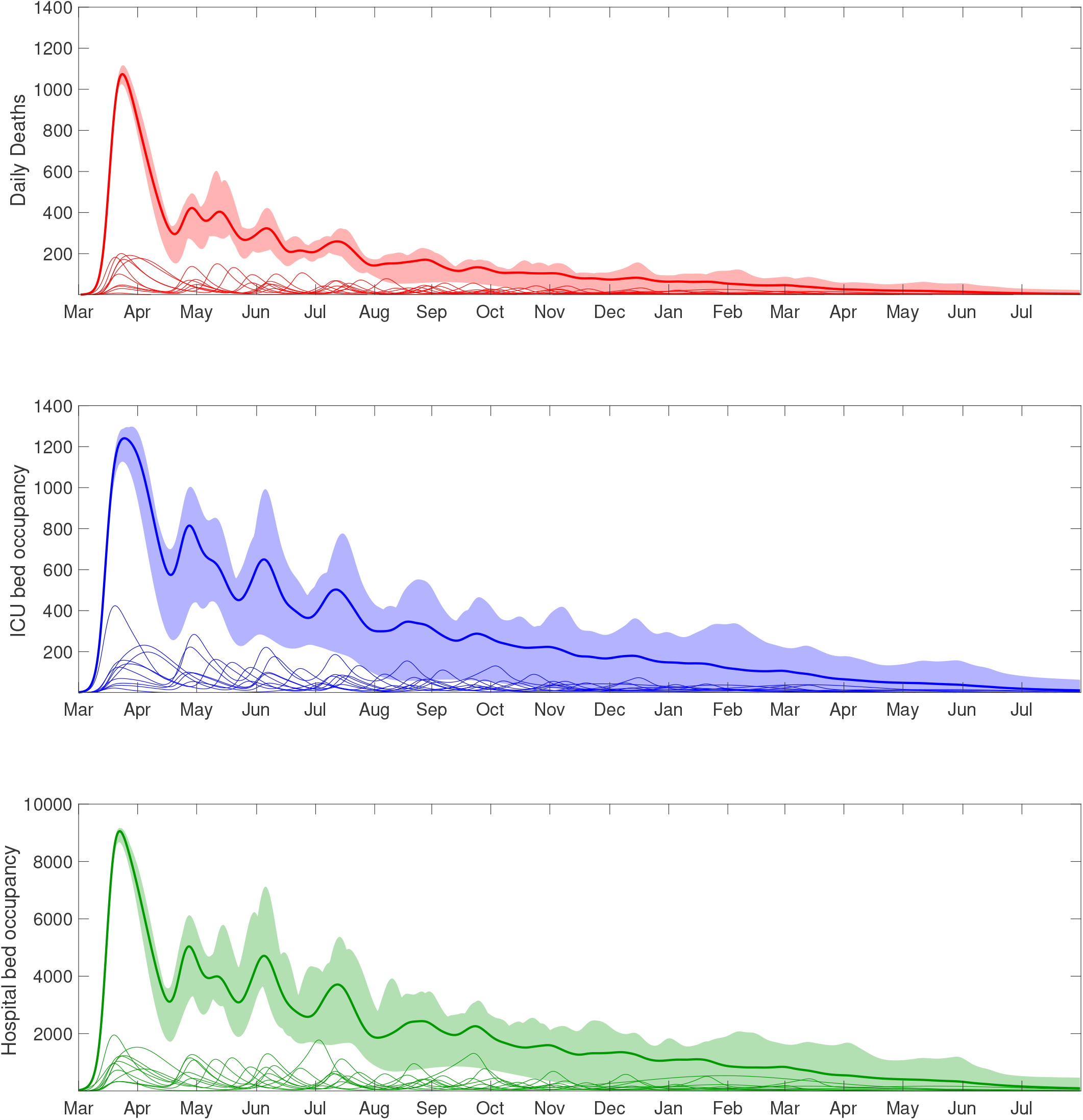
Clinical case projections under an adaptive intervention strategy with regionally activated lockdowns (responding to ICU occupancy). In all simulations we assumed social distancing measures were relaxed on 7th May for all individuals, with subsequent reintroduction of lockdown measures at a regional level (in the seven English regions, Scotland, Wales, and Northern Ireland) if ICU occupancy exceeded 45 ICU cases per million within the given region. Thick solid lines correspond to the mean outbreak over a sample of posterior parameters, with shaded regions corresponding to the 95% prediction intervals. The paler lines correspond to the dynamics in the individual regions. We display statistics on daily counts of **(Row one)** deaths; **(Row two)** ICU bed occupancy; **(Row three)** hospital bed occupancy.

### Role of asymptomatics crucial in determining the effect of age-based lockdown relaxation measures

Finally, we investigate the impact of applying lockdown measures for specific age groups, whilst varying *τ*, the relative level of transmission from asymptomatic individuals, and *α*, the scaling determining whether the age-dependence in cases comes from susceptibility (*α* = 1) or symptoms (*α* = 0). We observe that, regardless of the values of *τ* and *α*, applying control on only a single age group (PS, S, A or E) results in large-scale epidemics (Fig. 6). Similarly ineffective strategies are observed when combining PS control with one of S, A or E. However, control of school aged children, adults and the elderly, results in epidemics that are under control for all values of *τ* and *α*.

**Fig. 6:**
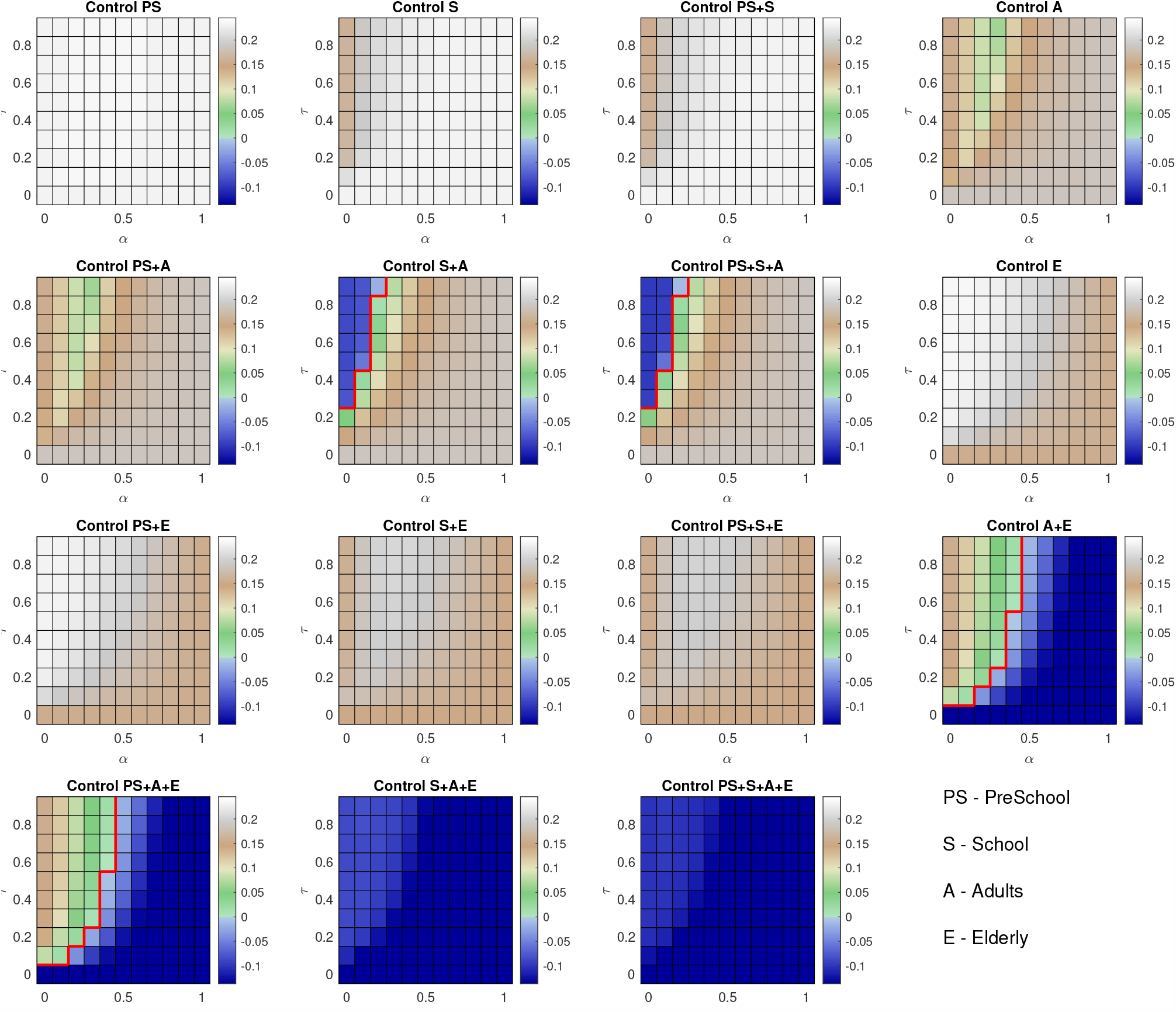
Sensitivity of intervention measures to *τ* and *α*. Each panel represents the application of lockdown measures to combinations of specific age groups (PS - PreSchool (0-4yrs), S - School (5-20yrs), A - Adults (21-70yrs), E - Elderly (over 70 yrs). The colour of each square represents the growth rate of the epidemic under the specified age-specific policies. Growth rates less than 0 (blue) imply that the epidemic is under control; the red line separates regions that are under control from regions where we expect exponential growth. Columns distinguish inclusion or exclusion of PS and S groupings in lockdown coverage: **(Column one)** coverage includes PS, not S; **(Column two)** coverage includes S, not PS; **(Column three)** coverage includes both PS and S; **(Column four)** neither PS or S included in lockdown.

Should we exempt the elderly from lockdown we find that, for high levels of *α*, large epidemics are observed, whereas if the true value of *τ* is high and *α* is small, applying control on the younger age groups and releasing lockdown on the elderly can result in epidemics that will rapidly die out. In contrast, if we relax lockdown on school children but keep it in place for other age groups, we note that this only has a positive effect upon the epidemic if the true value of *α* is high, or the true value of *τ* is low. If *α* is low and *τ* is high, then this implies that the age-dependence of reported cases is primarily as a result of clinical symptoms rather than susceptibility and the transmission rate of asymptomatic cases is high. Therefore, school children will play a much larger role in transmission, implying that a policy of re-opening school would cause a much larger epidemic. These results reinforce the need to resolve uncertainty regarding the role of asymptomatic individuals in the infection process in order to establish the optimal intervention strategy.

### Potential exit strategies comparison

Our findings are summarised in Table 2, where we focus on deaths (and the associated Quality Adjusted Life Year (QALY) losses), hospital occupancy and the scale of the lockdown as a measure of potential economic burden. QALYs are a standard measure in health economics which accounts for the number of life years lost due to an illness or disease, while also taking into account quality of life. Hence, under the QALY framework deaths in younger individuals have greater impact than deaths of older individuals due to the additional years of life lost (for further details, see the Supporting Text S5). Our lockdown scale measures the pro rata number of days the population is under lockdown; so if 50% of the population is under lockdown for 200 days, we report a value of 100 (50% of 200).

A completely uncontrolled outbreak is predicted to lead to around 200,000 deaths, approximately 2 million QALY losses but no lockdown impacts. If the current controls are maintained until the end of 2020, then we predict 39,000 deaths this year, but a further 159,000 if controls were then completely removed. Regional switching and age-dependent strategies provide alternative exit strategies in the absence of pharmaceutical interventions. Of these, the age-dependent shielding of those age 60 or over generates the lowest mortality and also the lowest lockdown scale, thereby minimising socio-economic disruption. However, it is unclear if a protracted lockdown of this age-group would be practical, ethical or politically acceptable.

## Discussion

In this paper, we have developed an age-structured compartmental SARS-CoV-2 transmission model that has been used to make short-term predictions and analyse the effectiveness of the strict social distancing measures that were implemented in the UK during April. The paper reflects the state of the model in April and our predictions at that time. We have not used the advantages of hindsight to improve the fits nor to change the scenarios considered [8]; instead, we use this discussion to consider what these results imply for the unfolding second wave and long-term exit strategies.

Our model shows that, without the introduction of the large scale social distancing measures that were introduced on 23rd March, the epidemic in the UK would have continued to grow exponentially and hospital and ICU occupancy would have rapidly exceeded capacity. However, under the enacted policies, the epidemic was predicted to peak in April for all regions of the UK, before starting to decline (Figure 2).

One of the most important questions postulated in April was when and how social distancing measures might be relaxed; a question that is still pertinent in late 2020. We consistently found that any relaxation of control measures in the short term leads to a rapid resurgence of COVID-19 disease with the health system potential being overwhelmed by a sizeable second epidemic wave (Fig. 3). In contrast, moderate or no adjustments to current social distancing measures allows hospital and ICU occupancy to remain within capacity over the duration of the outbreak, although this leaves dangerously high numbers of susceptible individuals in the population (Figure 3). It was apparent from the data on confirmed cases and deaths as a result of COVID-19 disease available in early April that the risks associated with infection increase with age [17–19]. We therefore also investigated the impact of age-specific control policies, whereby lockdown measures remained in place for all individuals over a certain age until the end of 2020 (Figure 4). We found that, whilst some marginal gains can be made should everyone over the age of 60 be put under isolation measures, extending this policy to include younger age groups increases the risk of a second wave occurring when measures are relaxed. Furthermore, we projected critical care to be stretched and ICU bed occupancy to exceed 4,000 during the course of the pandemic in all but the most wide-ranging age-specific lockdown policies (Fig. 4). This extreme form of shielding has since been advocated as a potential exit strategy [20, 21] but there are ethical issues as well as practical problems with isolating the most vulnerable from the rest of society.

Our sensitivity analysis shows that the effectiveness of any age-specific intervention policy is critically dependent upon the precise role of asymptomatic individuals in the epidemic. Even in April, undocumented infection has been inferred to have facilitated the spread of SARS-CoV-2 in China [22], suggesting the potential of asymptomatic transmission. At the time government advice for self-quarantining focused upon individuals who showed symptoms of COVID-19 (primarily a fever and a dry persistent cough) and therefore, our predictions for asymptomatic (or pre-symptomatic) infections playing a significant role in the transmission process, weaken such a policy. Asymptomatic transmission and transmission before symptom onset are now well recognised phenomena [23], which emphasises the need for efficient test-trace-and-isolate policies.

In practice, to minimise the risk of a second large epidemic wave occurring in the UK, adaptive policies may need to be considered that react to local health pressures. To that end, we examined a more bespoke intervention policy whereby measures were relaxed and re-introduced on a regional basis, with a defined trigger for the reintroduction of interventions when ICU occupancy exceeded a certain level. This results in a longer epidemic tail, until the second half of 2021, but ensures that the health service is protected by reintroducing social distancing measures for all individuals in a region when required.

In mid-April many countries around the world had now seen significant epidemics of COVID-19 and many had implemented severe lockdown policies in an effort to contain the disease. In China and other countries in East Asia, once the epidemic was regarded to be under control, in seeking to prevent the occurrence of a large second wave the relaxation of isolation measures was implemented in a gradual fashion, and was tightly reimposed if new cases were detected. Our model findings support the need for this form of relaxation policy. We recognise that there is a need for certain measures to be lifted as soon as is feasible, for a range of practical, social and economic reasons. However, government agencies should be prepared to resume lockdown if needed, based upon the the progression of the epidemic following relaxation. Identifying triggers, such as ICU occupancy exceeding a certain threshold, may be beneficial in allowing decision makers to follow a clear set of guidelines for controls to be reintroduced. The identification of such triggers need be based upon the objective of an intervention measure and the ability to resolve epidemiological uncertainty as the outbreak progresses. To this end, formal adaptive management approaches may help to facilitate the establishment of state dependent intervention strategies [24].

The model described is necessarily a simplified representation of reality based on several assumptions and has various limitations. Data informing contact structure for the UK were measured historically [13]. Were contact patterns in early 2020 (pre-lockdown) to substantially differ from the preexisting data, the influence of projected intervention effects may be impacted. Similarly, while we can infer the compliance to the currently imposed rules, we had limited understanding of how people would behave when the controls are released — would they remain wary of potentially infectious situations, or would they compensate for the time in lockdown. This still remains an open question [25] and is a key policy consideration as restrictions are varied. Throughout, we have assumed that when controls are lifted mixing patterns would return to their pre-pandemic norm.

Heterogeneities in compliance and in infection patterns, such as increased transmission in hospitals and institutions, may affect the outcome of the measures considered. We note that these early estimates of deaths resulting from an individual strategy does not take into account the potential for increased deaths due to exceeding hospital or ICU capacities, and so may underestimate deaths from strategies resulting in high occupancies. However, our April estimate of around 39,000 deaths from the first wave of COVID-19 infections in the UK compares well with the true figure of 41,265 from 1st August 2020 before cases began to rise again. In addition, though there have been recorded instances of superspreading events for COVID-19 [26], our model does not explicitly account for such highly stochastic dynamics. Such stochastic effects will be important at times of low infection (such as troughs between waves) and could influence the timing of a second wave. However, beyond the early stages of the outbreak the dynamics at the population-level are generally driven by the average pattern of social mixing, rather than individual level variation, meaning a deterministic framework is a justifiable approximation.

Since these results were produced in April, there have been multiple changes to the methodology precipitated largely by additional data, and the need to match to these new sources [16]. Three key additional data sets have shaped the model development. From mid-June age-structured data became available on antibody seroprevalence from weekly blood donor samples from different regions of England (approximately 1000 samples per region) [27]. Matching to serology allowed us to set an independent scaling between infections and epidemiological observations (such as hospitalisations and deaths), particularly important given that a significant proportion of infections are asymptomatic. The age-structured nature of this data also helps to refine the key parameter *α* in our model that determines the contribution of age-dependent susceptibility and age-dependent symptomatic probability. This has since been surpassed by serological data from the REACT2 study [**?**], which was a carefully designed sample of 100,000 individuals to gain a representative sample across England. Finally, community swab testing (through the Pillar 2 arm of the Test and Trace scheme), provides the most rapid assessment of current infection levels across the UK, without the delays associated with hospitalisation or death. Each of these requires a restructuring of the model framework to account for the new data stream. There have also been significant improvements in fitting the model to the data: integer quantities are assumed to be drawn from a negative binomial distribution, while proportions are drawn from a beta binomial — both leading to increased variance for a given mean. Finally, the impact of restrictions (*ϕ*) is allowed to vary slowly on a weekly time scale to account for the multiple changes in both policy and compliance.

All the strategies we have considered here assume that an exit strategy will have to rely on non-pharmaceutical interventions. In this case, a second (or subsequent) wave of infections follows any return to normality while there is sufficient susceptibility in the population. We are therefore faced with three potential exit solutions: 1) Seek a measured reduction in restrictions that minimises the impact of the unfolding outbreak, but acknowledging that a significant proportion of the population will become infected (although not necessarily symptomatic); 2) Accept a substantial and long-term change to our social interactions (practising far better prevention of transmission), such that the reproductive ratio of the virus is constantly held below one — electronic and traditional methods of tracing and isolation [28] fall into this category; or 3) rely on the development of an effective vaccine, in which case the best approach may be to extend the lockdown, reducing infection until mass vaccination can occur.

In conclusion, the COVID-19 pandemic has resulted in the introduction of multiple levels of social distancing measures in the UK and many other countries around the world. Following the strict lockdowns to mitigate the first wave, public-health agencies are continually analysing how best to develop an exit strategy that balances the epidemiological consequences against impacts on mental health and the economy. Our work provides strong evidence to support the need for a cautious, measured approach to relaxation of any controls, in order to provide necessary support for the health service and to protect the most vulnerable members of society.

## Data Availability

Data on cases were obtained from the COVID-19 Hospitalisation in England Surveillance System (CHESS) data set that collects detailed data on patients infected with COVID-19. Data on COVID-19 deaths were obtained from Public Health England. These data contain confidential information, with public data deposition non-permissible for socioeconomic reasons. The CHESS data resides with the National Health Service (www.nhs.gov.uk) whilst the death data are available from Public Health England (www.phe.gov.uk).

## Acknowledgements

The authors would like to thank the RAMP Rapid Review Group for their useful comments on this manuscript.

## Author contributions

**Conceptualisation:** Matt J. Keeling.

**Data curation:** Matt J. Keeling; Glen Guyver-Fletcher; Alexander Holmes, Massimiliano Tamborrino.

**Formal analysis:** Matt J. Keeling.

**Investigation:** Matt J. Keeling.

**Methodology:** Matt J. Keeling.

**Software:** Matt J. Keeling; Edward M. Hill; Louise Dyson; Michael J. Tildesley.

**Validation:** Matt J. Keeling; Edward M. Hill; Louise Dyson; Michael J. Tildesley.

**Visualisation:** Matt J. Keeling.

**Writing - original draft:** Michael J. Tildesley; Edward M. Hill; Louise Dyson; Matt J. Keeling.

**Writing - review & editing:** Matt J. Keeling; Edward M. Hill; Louise Dyson; Erin E. Gorsich; Bridget Penman; Trystan Leng; Glen Guyver-Fletcher; Alexander Holmes, Massimiliano Tamborrino; Hector McKimm; Michael J. Tildesley.

## Ethical considerations

The data were supplied from the CHESS database after anonymisation under strict data protection protocols agreed between the University of Warwick and Public Health England. The ethics of the use of these data for these purposes was agreed by Public Health England with the Government’s SPI-M(O) / SAGE committees.

## Supporting information items

**Supporting Text S1**

Description of the complete system of model equations.

**Supporting Text S2**

Information on the data streams informing public health measurable quantities.

**Supporting Text S3**

Distributions of key parameters from the MCMC process.

**Supporting Text S4**

Details on the mechanisms underpinning social distancing measures within the model framework.

**Supporting Text S5**

Explanation of the QALY losses computation.

**Table S1**

**EQ-5D index population norms for England**.

**Table S2**

**UK population aggregated to ten regions (rounded to nearest 10**,**000)**. With regard to our intervention scenario in which regional ICU occupancy triggered the reintroduction and relaxation of social distancing measures within that region, the final column lists each of the regional ICU bed occupancy thresholds (equating to 45 occupied ICU beds per one million population).

**Fig. S1**

**Key parameters inferred by the MCMC process**. The top two figures show the frequency distribution of *α* and *τ* which control the age-structured dynamics; the red line shows the uninformative prior ([0, 1] and [0.0.5] respectively. The middle row shows the results of the inferred *α* value, giving the distributions of *d*_*a*_ and *σ*_*a*_. The lower figure shows the impact of control measures *ϕ* in each of the ten regions. Throughout, error bars give the 95% credible interval, the box is the 50% credible interval and the line is the median value. (Predictions were produced on 23rd April, using data until 21st April).

**Fig. S2**

**Regional projections for hospital admissions per 100**,**000 with and without imposition of lockdown**. In each panel: filled markers correspond to observed data, solid lines correspond to the mean outbreak over a sample of posterior parameters; shaded regions depict prediction intervals, with darker shading representing stricter confidence (dark shading - 50%, moderate shading - 90%, light shading - 99%); dashed lines illustrate the mean projected trajectory had no lockdown measures being introduced (predictions were produced on 23rd April, using data until 21st April).

**Fig. S3**

**Regional projections for hospital occupancy per 100**,**000 with and without imposition of lockdown**. In each panel: filled markers correspond to observed data, solid lines correspond to the mean outbreak over a sample of posterior parameters; shaded regions depict prediction intervals, with darker shading representing stricter confidence (dark shading - 50%, moderate shading - 90%, light shading - 99%); dashed lines illustrate the mean projected trajectory had no lockdown measures being introduced (predictions were produced on 23rd April, using data until 21st April).

**Fig. S4**

**Regional projections for ICU bed occupancy per 100**,**000 with and without imposition of lockdown**. In each panel: filled markers correspond to observed data (squares are for reported deaths, circles are for death of death), solid lines correspond to the mean outbreak over a sample of posterior parameters; shaded regions depict prediction intervals, with darker shading representing stricter confidence (dark shading - 50%, moderate shading - 90%, light shading - 99%); dashed lines illustrate the mean projected trajectory had no lockdown measures being introduced.

