## Supplementary Information for "Predictions of COVID-19 dynamics in the UK: short-term forecasting and analysis of potential exit strategies"

#### Table of Contents

**1 Supporting Text S1: Model description** **2**

**2 Supporting Text S2: Public health measurable quantities** **5**

**3 Supporting Text S3: Key Inferred Parameters** **7**

**4 Supporting Text S4: Modelling social distancing** **8**

**5 Supporting Text S5: QALY losses** **9**

**6 Supporting Text S6: Additional tables** **10**

**7 Supporting Text S7: Additional figures** **11**

### 1 Supporting Text S1: Model description

We present here the system of equations that account for both household saturation of transmission and household quarantining. As described in the main text, individuals may be susceptible ($S$), exposed ($E$), with detectable infection ($D$), or undetectable infection (asymptomatic, $U$). Undetectable infections are assumed to transmit infection at a reduced rate given by $\tau$. We let superscripts denote the first infection in a household ($F$), a subsequent infection from a detectable/symptomatic household member ($SD$) and a subsequent infection from an asymptomatic household member ($SU$). A fraction ($H$) of the first detected case in a household is quarantined ($QF$), as are all their subsequent household infections ($QS$).

##### Model equations

The full equations are given by

$$
\begin{aligned}
\frac{dS_a}{dt} &= -\left(\lambda_a^F + \lambda_a^{SD} + \lambda_a^{SU} + \lambda_a^Q\right)\frac{S_a}{N_a},\\
\frac{dE_a^F}{dt} &= \lambda_a^F \frac{S_a}{N_a} - \epsilon E_a^F,\\
\frac{dE_a^{SD}}{dt} &= \lambda^{SD} \frac{S_a}{N_a} - \epsilon E_a^{SD},\\
\frac{dE_a^{SU}}{dt} &= \lambda^{SU} \frac{S_a}{N_a} - \epsilon E_a^{SU},\\
\frac{dE_a^Q}{dt} &= \lambda^Q S - \epsilon E_a^Q,\\
\frac{dD_a^F}{dt} &= d_a(1-H)\epsilon E_a^F - \gamma D_a^F,\\
\frac{dD_a^{SD}}{dt} &= d_a \epsilon E_a^{SD} - \gamma D_a^{SD},\\
\frac{dD_a^{SU}}{dt} &= d_a(1-H)\epsilon E_a^{SU} - \gamma D_a^{SU},\\
\frac{dD_a^{QF}}{dt} &= d_a H \epsilon E_a^F - \gamma D_a^{QF},\\
\frac{dD_a^{QS}}{dt} &= d_a H \epsilon E_a^{SU} + d_a \epsilon E_a^Q - \gamma D_a^{QS},\\
\frac{dU_a^F}{dt} &= (1-d_a)\epsilon E_a^F - \gamma U_a^F,\\
\frac{dU_a^S}{dt} &= (1-d_a)\epsilon (E_a^{SD} + E_a^{SU}) - \gamma U_a^S,\\
\frac{dU_a^Q}{dt} &= (1-d_a)\epsilon E_a^Q - \gamma U_a^Q,
\end{aligned}
$$

with the forces of infection obeying

$$\begin{aligned}
\lambda_a^F &= \sigma_a \sum_b \left(D_b^F + D_b^{SD} + D_b^{SU} + \tau(U_b^F + U_b^S)\right) \beta_{ba}^N, \\
\lambda_a^{SD} &= \sigma_a \sum_b D_b^F \beta_{ba}^H, \\
\lambda_a^{SU} &= \sigma_a \tau \sum_b U_a^F \beta_{ba}^H, \\
\lambda_a^Q &= \sigma_a \sum_b D_b^{QF} \beta_{ba}^H,
\end{aligned}$$

where $\beta_{ba}^H$ (with the subscript $ba$ corresponding to transmission from age group $b$ towards age group $a$) is household transmission and $\beta_{ba}^N = \beta_{ba}^S + \beta_{ba}^W + \beta_{ba}^O$ is all other transmission locations, comprising school-based transmission ($\beta_{ba}^S$), work-place transmission ($\beta_{ba}^W$) and transmission in all other locations ($\beta_{ba}^O$). $\sigma_a$ corresponds to the age-dependent susceptibility of individuals to infection, $d_a$ the age-dependent probability of displaying symptoms (and hence being detected), and $\tau$ represents reduced transmission of infection by undetectable individuals compared to detectable infections.

##### Amendments to within-household transmission

Given the novelty of the additional household structure that is included in this model, we clarify in more detail here the action of this formulation. We give a simpler set of equations (based on a standard $SIR$ model) that contains a similar household structure; in particular, we take the standard $SIR$ model and split the infected class into those first infected within a household ($I_F$) and subsequent infections ($I_S$):

$$\begin{aligned}
\frac{dS}{dt} &= -\beta^H S I_F - \beta^O S(I_F + I_S) \\
\frac{dI_F}{dt} &= \beta^O S(I_F + I_S) - \gamma I_F S \\
\frac{dI_S}{dt} &= \beta^H S I_F - \gamma I_S \\
\frac{dR}{dt} &= \gamma(I_F + I_S)
\end{aligned}$$

where the transmission rate is also split into within household transmission $\beta^H$ and all other transmission $\beta^O$ (i.e out-of-household transmission). Again, we make the assumption that only the first infection in any household generates infections within the household. We compare this to the SIR model without this additional structure:

$$\begin{aligned}
\frac{dS}{dt} &= -\widehat{\beta}^H SI - \widehat{\beta}^O SI \\
\frac{dI}{dt} &= \widehat{\beta}^H SI + \widehat{\beta}^O SI - \gamma I \\
\frac{dR}{dt} &= \gamma I
\end{aligned}$$

where we retain the split in transmission type.

The early growth rate of the two models are $\widehat{r} = \widehat{\beta}^H + \widehat{\beta}^O - \gamma$ for the simple SIR model, and $r = \frac{1}{2}\left[\beta^O - 2\gamma + \sqrt{\beta^{O2} + 4\beta^O\beta^H}\right]$ for the household structured version. From this simple comparison, it is clear that for the simple model the growth rate can remain positive even when control measures substantially reduce transmission outside the home ($\widehat{\beta}^O$ gets reduced), whereas in contrast for the structured version there is always a threshold level of transmission outside the household ($\beta_c^O = \gamma^2/(\beta^H + \gamma)$) that is needed to maintain positive growth.

For both the simple model given here and the full COVID-19 model, the inclusion of this additional household structure reduces the amount of within-household transmission compared to a model without this structure — as only the initial infection in each household ($I_F$) generates secondary within-household cases. It is therefore necessary to rescale the household transmission rate $\beta^H$ to obtain the appropriate average within-household attack rate. For the full COVID-19 model, we find that a simple multiplicative scaling to the household transmission ($\beta^H \to z\beta^H$, $z \approx 1.3$) generates a comparable match between the new model and a model without this household structure – even when age structure is included.

#### Relationship between age-dependent susceptibility and detectability

We interlink age-dependent susceptibility, $\sigma_a$, and detectability, $d_a$, by a quantity $Q_a$. $Q_a$ can be viewed as the scaling between force of infection and symptomatic infection. Taking a next-generation approach, the early dynamics would be specified by:

$$R_0 D_a = d_a \sigma_a \beta^N_{ba} \left(D_a + \tau U_a\right)/\gamma \qquad R_0 U_a = (1-d_a)\sigma_a \beta^N_{ba} \left(D_a + \tau U_a\right)/\gamma$$

where $D_a$ measures those with detectable infections, which mirrors the early recorded age distribution of symptomatic cases. Explicitly, we let $d_a = \frac{1}{\kappa} Q_a^{(1-\alpha)}$ and $\sigma_a = \frac{1}{k} Q_a^{\alpha}$. As a consequence, $Q_a = \kappa k d_a \sigma_a$; where the parameters $\kappa$ and $k$ are determined such that the oldest age groups have a 90% probability of being symptomatic ($d_{>90} = 0.90$) and such that the basic reproductive ratio from these calculations gives $R_0 = 2.7$.

#### 2 Supporting Text S2: Public health measurable quantities

The main model equations focus on the epidemiological dynamics, allowing us to compute the number of symptomatic and asymptomatic infectious individuals over time. However, these quantities are not measured - and even the number of confirmed cases (the closest measure to symptomatic infections) is highly biased by the testing protocols at any given point in time. It is therefore necessary to convert infection estimates into quantities of interest that can be compared to data. We considered five such quantities which we calculated from the number of newly detectable symptomatic infections on a given day $nD_d$.

1. **Hospital Admissions:** We assume that a fraction $P_a^{D\to H}$ of detectable cases will be admitted into hospital after a delay $q$ from the onset of symptoms. The delay, $q$, is drawn from a distribution $D_q^{D\to H}$ (note that $\sum_q D_q^{D\to H} = 1$.) Hospital admissions on day $d$ of age $a$ are therefore given by
$$H_a(d) = P_a^{D\to H} \sum_q D_q^{D\to H} nD_{d-q}$$

2. **ICU Admissions:** Similarly, a fraction $P_a^{D\to I}$ of detectable cases will be admitted into ICU after a delay, drawn from a distribution $D_q^{D\to I}$ which determines the time between the onset of symptoms and admission to ICU. ICU admissions on day $d$ of age $a$ are therefore given by
$$ICU_a(d) = P_a^{D\to I} \sum_q D_q^{D\to I} nD_{d-q}$$

3. **Hospital Beds Occupied:** Individuals admitted to hospital spend a variable number of days in hospital. We therefore define two weightings, which determine if someone admitted to hospital still occupies a hospital bed $q$ days later ($T_q^H$) and if someone admitted to ICU occupies a hospital bed on a normal ward $q$ days later ($T_q^{I\to H}$). Hospital beds occupied on day $d$ of age $a$ are therefore given by
$$H_a^o(d) = \sum_q H_a(d-q)T_q^H + \sum_q ICU_a(d-q)T_q^{I\to H}$$

4. **ICU Beds Occupied:** We similarly define $T_q^I$ as the probability that someone admitted to ICU is still occupying a bed in ICU $q$ days later. ICU beds occupied on day $d$ of age $a$ are therefore given by
$$ICU_a^o(d) = \sum_q ICU_a(d-q)T_q^I$$

5. **Number of Deaths:** The mortality ratio $P_a^{H\to Death}$ determines the probability that a hospitalised case of a given age, $a$, dies after a delay, $q$ drawn from a distribution, $D_d^{H\to Death}$ between hospitalisation and death. The number of deaths on day $d$ of age $a$ are therefore given by
$$\text{Deaths}_a(d) = P_a^{H\to Death} \sum_q H_a(d-q) D_d^{H\to Death}$$

These nine distributions are all parameterised from individual patient data as recorded by the COVID-19 Hospitalisation in England Surveillance System (CHESS) [1].

However, these distributions all represent a national average and do not therefore reflect regional differences. We therefore define regional scalings of the three key probabilities ($P_a^{D\to H}$, $P_a^{D\to I}$ and

$P_a^{H \to Death}$) and two additional parameters that can stretch (or contract) the distribution of times spent in hospital and ICU. These five regional parameters are necessary to get good agreement between key observations in all regions and may reflect both differences in risk groups (in addition to age) between regions or differences in how the data are recorded between devolved nations. We stress that these parameters do not (of themselves) influence the epidemiological dynamics, but do strongly influence how we fit to the evolving dynamics.

### 3 Supporting Text S3: Key Inferred Parameters

We show the distribution of key parameters from the MCMC process in Figure S1. We predict a moderate $\alpha$ and low $\tau$; the moderate $\alpha$ translates into a sharp change in susceptibility with age ($\sigma_a$) but a more steady change in the probability of symptoms. We also show the impact of restrictions ($\phi$) for the ten regions within the model.

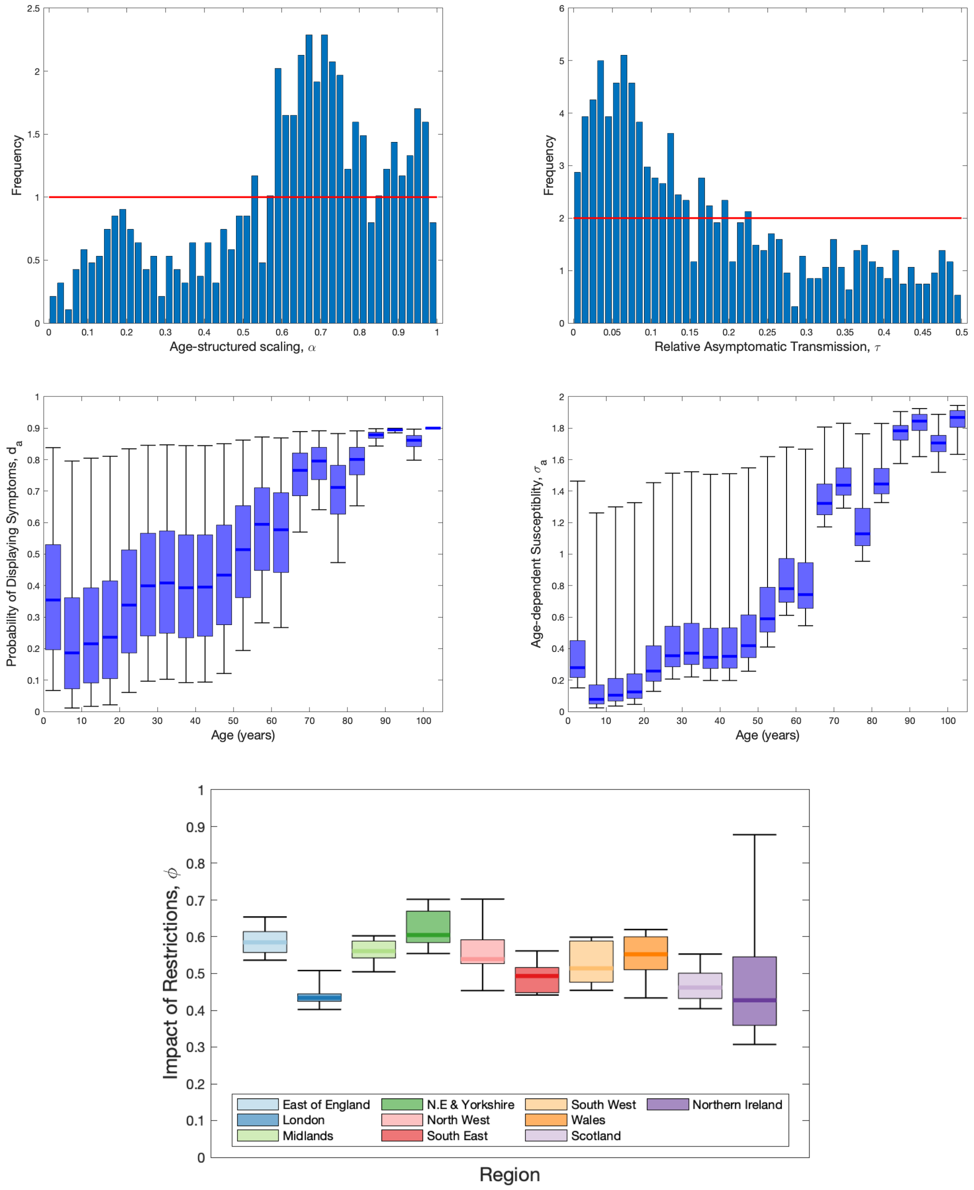


**Fig. S1: Key parameters inferred by the MCMC process.** The top two figures show the frequency distribution of $\alpha$ and $\tau$ which control the age-structured dynamics; the red line shows the uninformative prior ($[0, 1]$ and $[0.0.5]$ respectively. The middle row shows the results of the inferred $\alpha$ value, giving the distributions of $d_a$ and $\sigma_a$. The lower figure shows the impact of control measures $\phi$ in each of the ten regions. Throughout, error bars give the 95% credible interval, the box is the 50% credible interval and the line is the median value. (Predictions were produced on 23rd April, using data until 21st April).

### 4 Supporting Text S4: Modelling social distancing

Age-structured contact matrices for the United Kingdom were obtained from Prem et al. [2] and used to provide information on household transmission ($\beta^H_{ba}$, with the subscript $ba$ corresponding to transmission from age group $b$ towards age group $a$), school-based transmission ($\beta^S_{ba}$), work-place transmission ($\beta^W_{ba}$) and transmission in all other locations ($\beta^O_{ba}$). We assumed that the suite of social-distancing and lockdown measured acted in concert to reduce the work, school and other matrices while increasing the strength of household contacts.

We capture the impact of social-distancing by defining new transmission matrices ($B_{ba}$) that represent the potential transmission in the presence of extreme lockdown. In particular, we assume that:

$$B^S_{ba} = q^S \beta^S_{ba}, \qquad B^W_{ba} = q^W \beta^W_{ba}, \qquad B^O_{ba} = q^O \beta^O_{ba},$$

while household mixing $B^H$ is increased by up to a quarter to account for the greater time spent at home. We take $q^S = 0.05$, $q^W = 0.2$ and $q^O = 0.05$ to approximate the reduction in attendance at school, attendance at workplaces and engagement with shopping and leisure activities during the lock-down, respectively.

For a given compliance level, $\phi$, we generate new transmission matrices as follows:

$$\begin{aligned}
\widehat{\beta}^H_{ba} &= (1-\phi)\beta^H_{ba} + \phi B^H_{ba} \\
\widehat{\beta}^S_{ba} &= (1-\phi)\beta^S_{ba} + \phi B^S_{ba} \\
\widehat{\beta}^W_{ba} &= (1-\theta)\left[(1-\phi)\beta^W_{ba} + \phi B^W_{ba}\right] + \theta\left((1-\phi) + \phi q^W\right)((1-\phi) + \phi q^O)\beta^W_{ba} \\
\widehat{\beta}^O_{ba} &= \beta^O_{ba}((1-\phi) + \phi q^O)^2
\end{aligned}$$

As such, home and school interactions are scaled between their pre-lockdown values ($\beta$) and post-lockdown limits ($B$) by the scaling parameter $\phi$. Work interactions that are not in public-facing 'industries' (a proportion 1 - $\theta$) were also assumed to scale in this manner; while those that interact with the general populations (such as shop-workers) were assumed to scale as both a function of their reduction and the reduction of others. We have assumed $\theta = 0.3$ throughout. Similarly, the reduction in transmission in other settings (generally shopping and leisure) has been assumed to scale with the reduction in activity of both members of any interaction, giving rise to a squared term.

### 5 Supporting Text S5: QALY losses

The loss of quality adjusted life years (QALYs) due to deaths was based on the quality adjusted life expectancy by age, and modified for the relative life-expectancy of individuals that die:

$$\text{Fatal case QALY loss} = \sum_{a=1}^{21} \left(D(a) \times E(a)\right),$$

whereby $D(a)$ is the number of deaths in age bracket $a$, and $E(a)$ is the discounted quality adjusted value of the remaining life expectancy, $L(a)$, of individuals in age group $a$. This quality adjusted life expectancy is given by:

$$E(a) = \sum_{i=1}^{L(a)} \frac{Q_w(\hat{a} + i)}{(1+d)^i}$$

where $Q_w(a)$ is the age-specific quality of life weight at age $a$, $\hat{a}$ is the average age (in years) of an individual in age-group $a$, and $d$ the discount rate (set at 0.035, corresponding to 3.5% per annum); the values of $L(a)$ are rounded to full years.

For parameterising the age-specific quality of life weights, $Q_w$, we obtained age-specific EQ-5D index population norms estimates for England from two literature sources. We took childhood estimates (which we used to cover 0–19 years of age) from Table 3 of Kwon *et al.* [3], and values for those aged 20 and above were sourced from Table 3.6 of Janssen and Szende [4]. A complete listing of age-specific quality of life weights values by age is presented in Table S1.

**Table S1: EQ-5D index population norms for England.**

| Age group (yrs) | EQ-5D index population norms scale |
|---|---|
| <20 | 0.948 |
| 20–24 | 0.929 |
| 25–34 | 0.919 |
| 35–44 | 0.893 |
| 45–54 | 0.855 |
| 55–64 | 0.810 |
| 65–74 | 0.773 |
| 75+ | 0.703 |

### 6 Supporting Text S6: Additional tables

**Table S2: UK population aggregated to ten regions (rounded to nearest 10,000).** For our intervention scenario in which regional ICU occupancy triggered the reintroduction and relaxation of social distancing measures within that region, the final column lists each of the regional ICU bed occupancy thresholds (equating to 45 occupied ICU beds per one million population).

| **Region** | **Population (millions)** | **Intervention trigger threshold (ICU bed occupancy)** |
|---|---:|---:|
| Wales | 3.14 | 142 |
| Scotland | 5.44 | 245 |
| Northern Ireland | 1.88 | 85 |
| East of England | 6.20 | 280 |
| London | 8.91 | 401 |
| Midlands | 10.70 | 482 |
| North East and Yorkshire | 8.14 | 367 |
| North West England | 7.29 | 328 |
| South East England | 9.13 | 412 |
| South West England | 5.60 | 252 |

### 7 Supporting Text S7: Additional figures

Here we show the model fit to the available data on hospital bed occupancy, hospital admissions and ICU bed occupancy, which are combined with the data on deaths (see main text figure 2). It is clear that due to their greater number fitting to hospital occupancy dominates the fitting process.

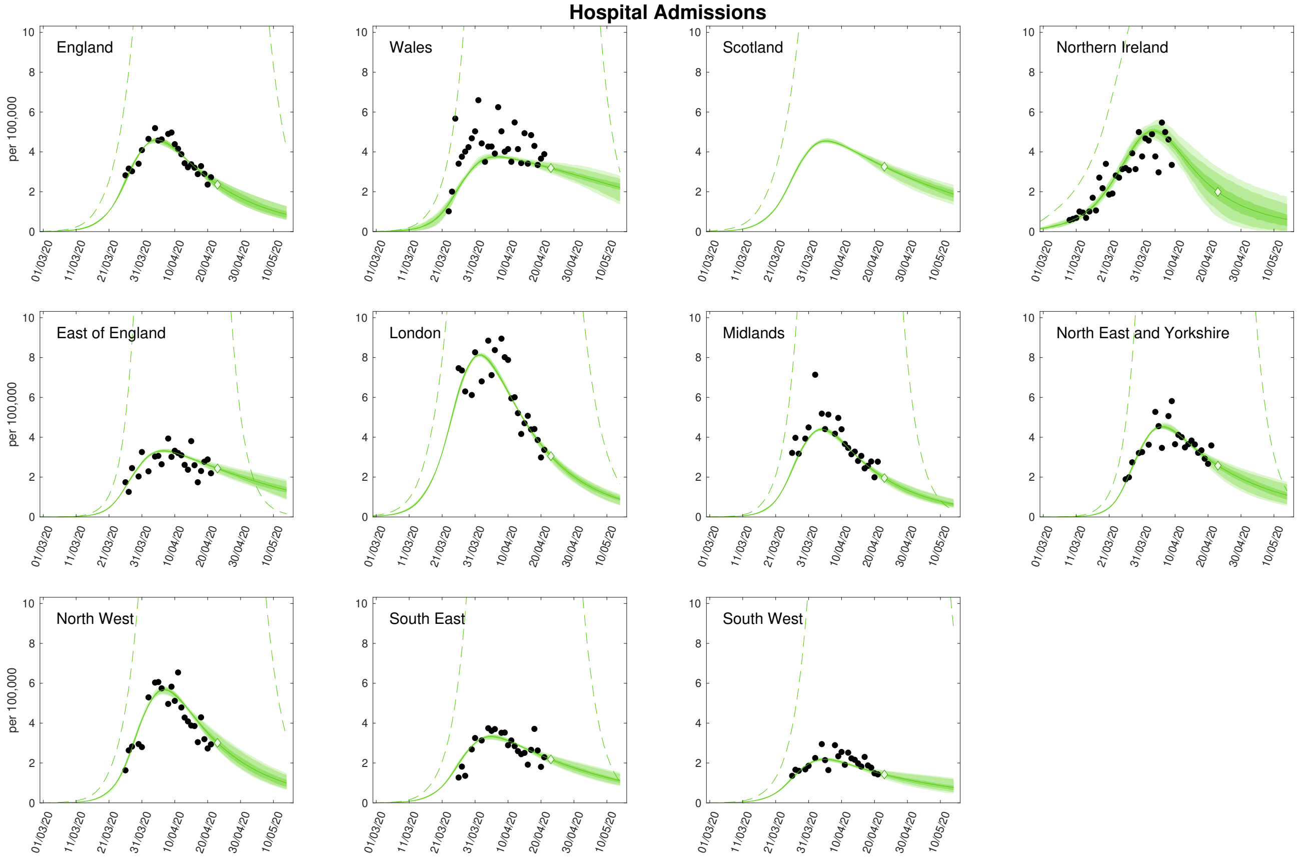


**Fig. S2: Regional projections for hospital admissions per 100,000 with and without imposition of lockdown.** In each panel: filled markers correspond to observed data, solid lines correspond to the mean outbreak over a sample of posterior parameters; shaded regions depict prediction intervals, with darker shading representing stricter confidence (dark shading - 50%, moderate shading - 90%, light shading - 99%); dashed lines illustrate the mean projected trajectory had no lockdown measures being introduced (predictions were produced on 23rd April, using data until 21st April).

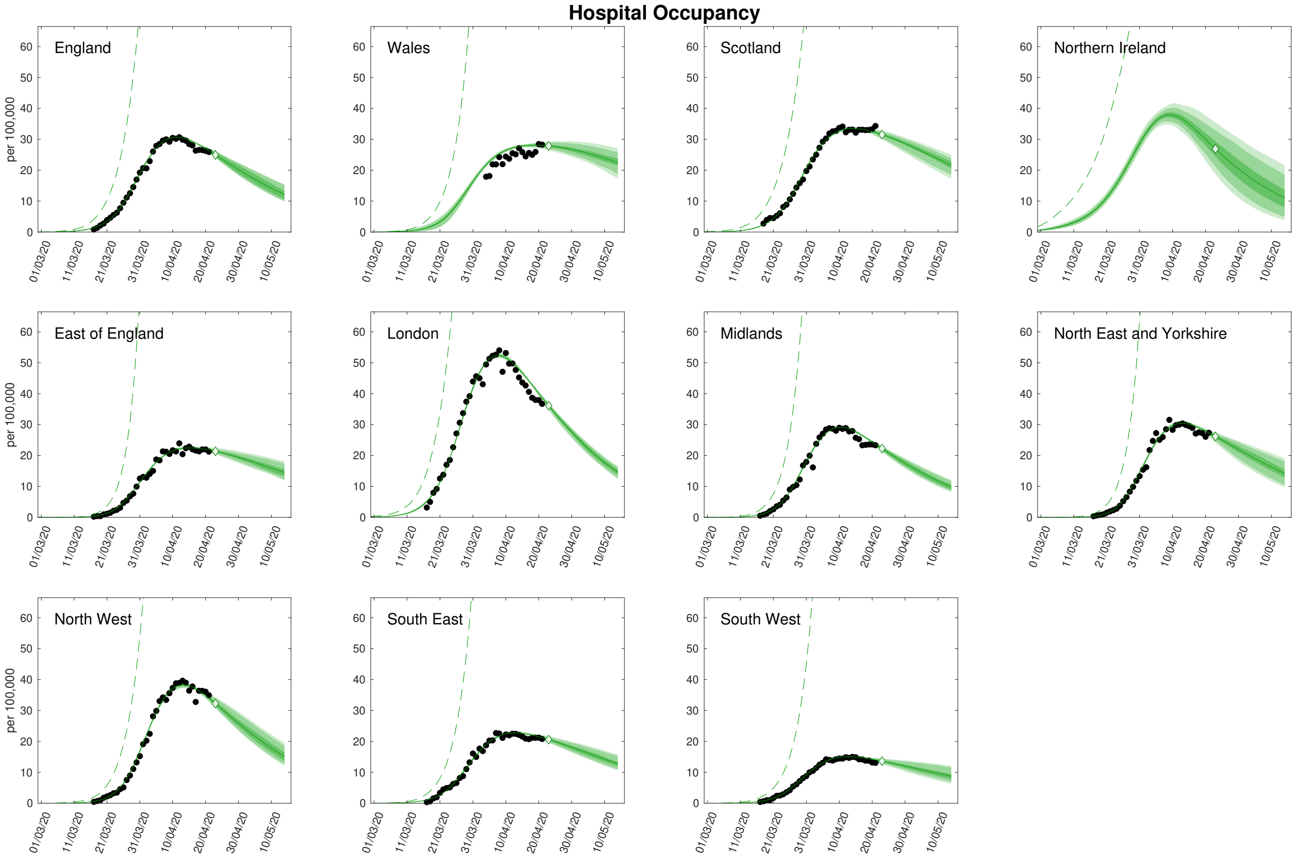


**Fig. S3: Regional projections for hospital occupancy per 100,000 with and without imposition of lockdown.** In each panel: filled markers correspond to observed data, solid lines correspond to the mean outbreak over a sample of posterior parameters; shaded regions depict prediction intervals, with darker shading representing stricter confidence (dark shading - 50%, moderate shading - 90%, light shading - 99%); dashed lines illustrate the mean projected trajectory had no lockdown measures being introduced (predictions were produced on 23rd April, using data until 21st April).

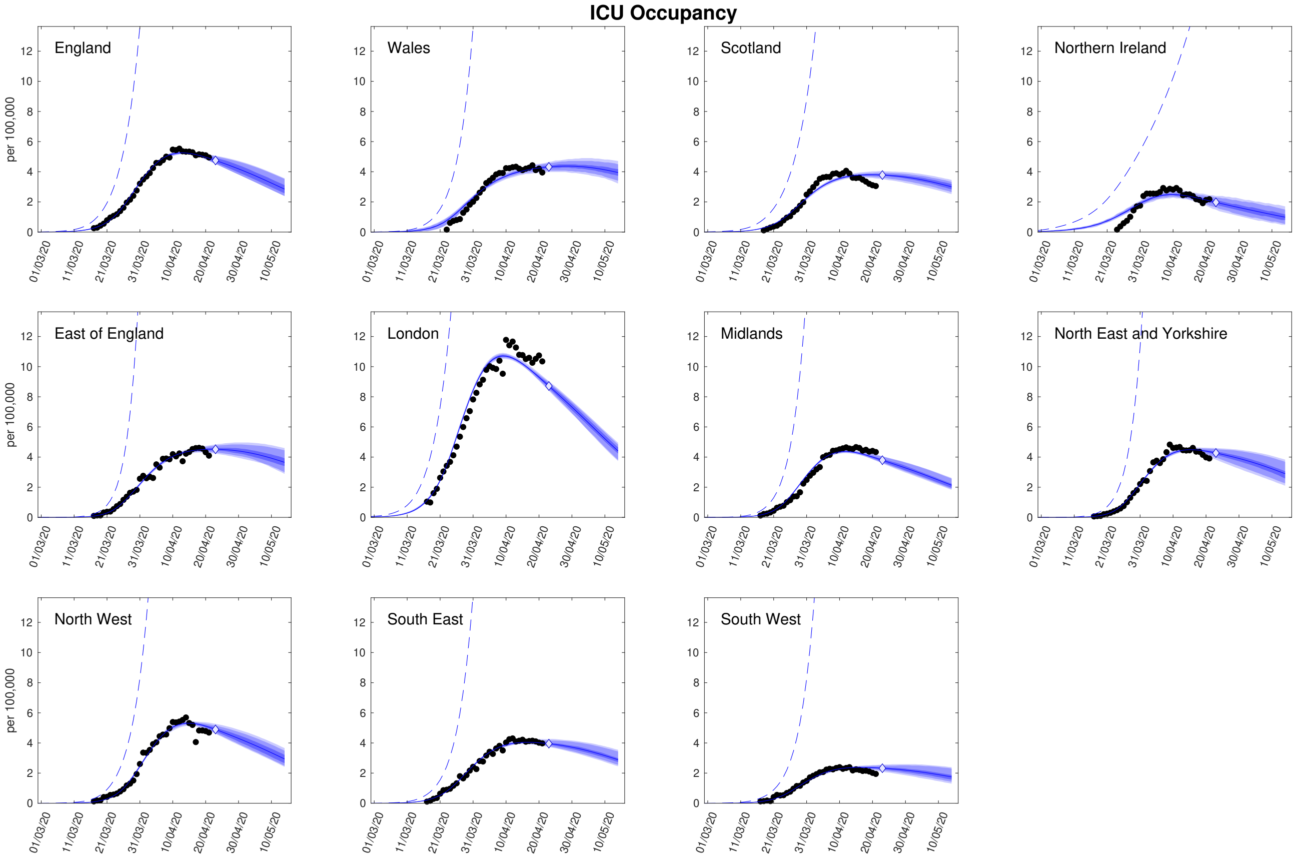


**Fig. S4: Regional projections for ICU bed occupancy per 100,000 with and without imposition of lockdown.** In each panel: filled markers correspond to observed data, solid lines correspond to the mean outbreak over a sample of posterior parameters; shaded regions depict prediction intervals, with darker shading representing stricter confidence (dark shading - 50%, moderate shading - 90%, light shading - 99%); dashed lines illustrate the mean projected trajectory had no lockdown measures being introduced (predictions were produced on 23rd April, using data until 21st April).
